# The impact of demographic factors on numbers of COVID-19 cases and deaths in Europe and the regions of Ukraine

**DOI:** 10.1101/2022.01.05.22268787

**Authors:** Igor Nesteruk, Oleksii Rodionov

## Abstract

The accumulated numbers of COVID-19 cases and deaths per capita are important characteristics of the pandemic dynamics that may also indicate the effectiveness of quarantine, testing, vaccination, and treatment. The statistical analysis based on the number of cases per capita accumulated to the end of June 2021 showed no correlations with the volume of population, its density, and the urbanization level both in European countries and regions of Ukraine. The same result was obtained with the use of fresher datasets (as of December 23, 2021). The number of deaths per capita and per case may depend on the urbanization level. For European countries these relative characteristics decrease with the increase of the urbanization level. Opposite trend was revealed for the number of deaths per capita in Ukrainian regions.

## Introduction

The accumulated number of COVID-19 cases per capita (CC) and deaths per capita (DC) may indicate the effectiveness of quarantine, testing, vaccination, treatment, and also characterizes the virulence of coronavirus strains which circulated in a particular region. An important characteristic of the strain virulence and treatment efficiencies can be the number of deaths per case DC/CC. The CC and DC numbers are regularly reported by World Health Organization, [1] and COVID-19 Data Repository by the Center for Systems Science and Engineering (CSSE) at Johns Hopkins University (JHU), [2].

The impact of some eco-demographic factors on the COVID-19 pandemic dynamics was studied in [3-20]. In particular, in [20] the influences of the volume of population, its density, and the urbanization level on the CC numbers accumulated at the end of June 2021 were investigated for European countries and the regions of Ukraine. Since no statistically significant correlations were revealed, in this paper we will study the influence of the volume of population *N*_*pop*_, its density, and the level of urbanization *N*_*ubr*_/*N*_*pop*_ (*N*_*ubr*_ is the number of people living in cities) on the CC, DC and DC/DC values with the use of fresher figures (as of December 23, 2021) for European countries and regions of Ukraine.

### Data

For this statistical analysis we will use the data set regarding the numbers of laboratory-confirmed COVID-19 cases in the regions of Ukraine accumulated at the time December 23, 2021 and compare it with the results based on the figures accumulated at June 27, 2021. As in paper [20], we will use the CC numbers (per 100 persons of population) registered by national statistics [21] and demographic data sets for Ukrainian regions [22] (see Table 1). The accumulated numbers of deaths registered by national statistics [21] in Ukrainian regions at December 23, 2021 are shown in the last column of Table 1. As the information from the regions of Ukraine fully or partially occupied by the Russian Federation is inaccurate, we excluded from consideration Donetsk and Luhansk regions, Crimea and Sevastopol. The CC figures (per 1,000,000 persons of population) registered by JHU [2] at two moments of time: June 28, 2021 and December 23, 2021 are shown in Table 2, which contains also the accumulated number of deaths per million (DC) as of December 23, 2021, [2] and the demographic data sets for European countries taken from [23-25].

**Table 1.** Demographic characteristics and the accumulated number of laboratory-confirmed COVID-19 cases and deaths in the regions of Ukraine as of June 27, 2021 and December 23, 2021.

| Oblast (region) | Population<br>$N_{pop}$,<br>[22] | Urban popu-<br>lation,<br>$N_{ubr}$,<br>[22] | Popula-<br>tion<br>density<br>(number<br>of<br>people<br>per<br>square<br>km), [22] | Accumu-<br>lated<br>number of<br>laboratory-confirmed<br>cases per<br>hundred as<br>of June 27,<br>2021, [21] | Accumu-<br>lated<br>number of<br>laboratory-confirmed<br>cases per<br>hundred as<br>of Decem-<br>ber 23,<br>2021, [21] | Accumu-<br>lated<br>number<br>of deaths<br>as of Decem-<br>ber 23,<br>2021,<br>[21] |
| --- | --- | --- | --- | --- | --- | --- |
| Vinnytsia | 1 545 416 | 799 385 | 58.29 | 4.67 | 8.8 | 3029 |
| Volyn | 1 031 421 | 539 179 | 51.2 | 6.06 | 10.03 | 2148 |
| Dnipropetrovsk | 3 176 648 | 2 668 744 | 99.54 | 4.33 | 7.75 | 8707 |
| Zhytomyr | 1 208 212 | 716 457 | 40.5 | 7.43 | 11.79 | 3204 |
| Zakarpattia | 1 253 791 | 465 904 | 98.13 | 4.98 | 6.96 | 2283 |
| Zaporizhzhia | 1 687 401 | 1 306 231 | 62.08 | 6.30 | 10.99 | 5105 |
| Ivano-Frankivsk | 1 368 097 | 606 764 | 98.23 | 6.38 | 9.32 | 3052 |
| Kyiv (oblast) | 1 781 044 | 1 105 383 | 63.31 | 7.12 | 10.21 | 4588 |
| Kirovohrad | 933 109 | 591 944 | 37.95 | 2.25 | 3.59 | 1458 |
| Lviv | 2 512 084 | 1 534 040 | 115.06 | 5.52 | 8.78 | 5900 |
| Mykolaiv | 1 119 862 | 768 022 | 45.53 | 6.34 | 10.45 | 3231 |
| Odesa | 2 377 230 | 1 597 062 | 71.37 | 5.95 | 10.1 | 5488 |
| Poltava | 1 386 978 | 867 201 | 48.25 | 5.70 | 10.16 | 3619 |
| Rivne | 1 152 961 | 548 088 | 57.51 | 6.93 | 11.07 | 2231 |
| Sumy | 1 068 247 | 741 430 | 44.82 | 7.43 | 12.5 | 2781 |
| Ternopil | 1 038 695 | 473 727 | 75.14 | 6.82 | 10.4 | 2075 |
| Kharkiv | 2 658 461 | 2 158 121 | 84.62 | 5.66 | 9.27 | 6231 |
| Kherson | 1 027 913 | 631 317 | 36.12 | 3.53 | 7.81 | 2656 |
| Khmelnyskyi | 1 254 702 | 720 752 | 60.82 | 7.18 | 11.78 | 3262 |
| Cherkasy | 1 192 137 | 678 682 | 57.04 | 6.97 | 10.92 | 2526 |
| Chernivtsi | 901 632 | 390 551 | 111.35 | 8.94 | 13.3 | 2976 |
| Chernihiv | 991 294 | 649 063 | 31.11 | 5.92 | 9.74 | 2353 |
| Kyiv (city) | 2 967 360 | 2 967 360 | 3536.78 | 7.22 | 11.01 | 8069 |

**Table 2.** Demographic characteristics and the accumulated number of laboratory-confirmed COVID-19 cases and deaths in European countries as of June 28, 2021 and December 23, 2021.

| Country | Population, [23] | Urbanization level $N_{ubr}/N_{pop}$ %, [24] | Population density (per square km), [25] | Total cases per million CC as of June 28, 2021, [2] | Total cases per million CC as of December 23, 2021, [2] | Total deaths per million DC as of December 23, 2021, [2] |
| --- | --- | --- | --- | --- | --- | --- |
| Monaco | 38682 | 100 | 1,547 | 65615.126 | 116548.6 | 961.538 |
| Vatican City | 801 | 100 | 2,273 | 33374.536 | 33251.23 | no data |
| Malta | 514564 | 95 | 1,447 | 69330.229 | 84878.9 | 916.489 |
| San Marino | 33785 | 97 | 561 | 150008.84 | 218965 | 2852.102 |
| Netherlands | 17059560 | 91 | 416 | 99886.646 | 176606.8 | 1205.141 |
| Belgium | 11482178 | 98 | 384 | 93486.963 | 174589 | 2416.54 |
| United Kingdom | 67141684 | 83 | 270 | 70284.988 | 172901.5 | 2167.399 |
| Liechtenstein | 37910 | 14 | 245 | 79555.288 | 154127.7 | 1803.733 |
| Luxembourg | 604245 | 91 | 243 | 112850.972 | 155069 | 1428.765 |
| Germany | 83124418 | 77 | 225 | 44576.917 | 83172 | 1312.555 |
| Italy | 60627291 | 70 | 207 | 70432.141 | 91391.17 | 2256.927 |
| Switzerland | 8525611 | 74 | 204 | 81198.962 | 140829.1 | 1383.398 |
| Andorra | 77006 | 88 | 183 | 179667.379 | 278860.8 | 1796.934 |
| Denmark | 5752126 | 88 | 136 | 50743.387 | 115907.8 | 545.817 |
| Czech Republic | 10665677 | 74 | 136 | 155658.773 | 224437.2 | 3290.394 |
| Poland | 37921592 | 60 | 122 | 76088.436 | 106289.4 | 2472.286 |
| Portugal | 10256193 | 65 | 112 | 85856.051 | 123239.9 | 1852.886 |
| Slovakia | 5453014 | 54 | 111 | 71720.074 | 245814.2 | 2973.78 |
| Albania | 2882740 | 60 | 107 | 46046.633 | 72029.15 | 1107.23 |
| Austria | 8891388 | 66 | 106 | 72206.875 | 139153.7 | 1503.361 |
| France | 64990511 | 80 | 105 | 86325.089 | 132236.8 | 1810.972 |
| Hungary | 9707499 | 71 | 105 | 83645.21 | 128431.5 | 3976.163 |
| Turkey | 82340088 | 75 | 105 | 64196.94 | 108763.9 | 953.932 |
| Slovenia | 2077837 | 55 | 104 | 123742.383 | 217967 | 2659.325 |
| Moldova | 4051944 | 43 | 99 | 63613.375 | 92880.14 | 2379.458 |
| Spain | 46692858 | 80 | 99 | 81117.733 | 122322.8 | 1904.345 |
| Serbia | 8802754 | 56 | 91 | 105279.579 | 187323.5 | 1821.715 |
| Romania | 19506114 | 54 | 89 | 56174.491 | 94160.52 | 3056.446 |
| North Macedonia | 2082957 | 58 | 83 | 74722.806 | 106754.3 | 3777.859 |
| Greece | 10522246 | 79 | 80 | 40416.745 | 101881.1 | 1947.594 |
| Bosnia and Herzegovina | 3323925 | 48 | 75 | 62481.731 | 87879.76 | 4057.964 |
| Croatia | 4156405 | 57 | 75 | 87610.845 | 168213.5 | 2983.837 |
| Ireland | 4818690 | 63 | 74 | 55002.07 | 136541.9 | 1182.042 |
| Ukraine | 44246156 | 69 | 73 | 52556.15 | 87621.13 | 2304.263 |
| Bulgaria | 7051608 | 75 | 63 | 60682.066 | 106153.8 | 4413.154 |
| Belarus | 9452617 | 78 | 46 | 44001.045 | 72981.12 | 577.049 |
| Montenegro | 627809 | 67 | 44 | 159544.758 | 257851.7 | 3802.239 |
| Lithuania | 2801264 | 68 | 42 | 102372.965 | 188826 | 2685.268 |
| Latvia | 1928459 | 68 | 29 | 72759.97 | 144706.8 | 2404.477 |
| Estonia | 1322920 | 69 | 27 | 98740.406 | 177301.6 | 1433.004 |
| Sweden | 9971638 | 87 | 23 | 107819.278 | 125324.1 | 1502.437 |
| Norway | 5337962 | 82 | 17 | 24116.983 | 67305.34 | 229.983 |
| Finland | 5522576 | 85 | 16 | 17176.113 | 40943.44 | 271.071 |
| Iceland | 336713 | 94 | 3 | 19208.791 | 64326.07 | 107.759 |

### Linear regression and Fisher tests

The linear regression will be used to calculate the regression coefficient *r* and the coefficients *a* and *b* of corresponding straight lines, [26]:

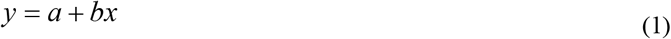

where *x* is the volume of population *N*_*pop*_, its density per square km, or the urbanization level N_urb_/*N*_*pop*_; *y* is the numbers of cases per capita CC, numbers of deaths per capita DC and deaths per case ratio DC/CC.

We will also use the F-test for the null hypothesis that says that the proposed linear relationship (1) fits the data sets. The experimental values of the Fisher function can be calculated with the use of the formula:

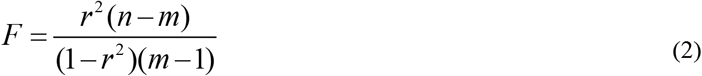

where *m*=2 is the number of parameters in the regression equation, [26]. The experimental values *F* must be compared with the critical values *F*_*C*_ (*k*_1_, *k*_2_) of the Fisher function at a desired significance or confidence level (*k*_1_ = *m* -1, *k*_2_ = *n* - *m*, see, e.g., [27]). If *F*_*C*_ (*k*_1_, *k*_2_) > *F*, the corresponding hypothesis is supported by observations.

## Results

Within six months, the numbers of cases per hundred in the regions of Ukraine increased from 1.4 to 2.2 times (see Table 1 and Fig. 1). The minimum values remained in the Kirovohrad region 3.59 (2.25), and the maximum ones - in the Chernivtsi region 13.3 (8.94). The variation (the difference between maximum and minimum values) increased 1.45 times. Rather high CC values still remained in Zhytomyr, Khmelnytskyi, Kyiv (city), and Sumy regions. As of December 23, 2021 the maximum values of deaths per 100,000 persons were registered in Chernivtsi (330.1) and Zaporizhzhia (302.5) regions. The minimum values corresponding to Kirovohrad (156.3) and Zakarpattia (182.1) regions are approximately twice smaller (see Table 1). The numbers of deaths per 100 registered cases vary much more: from 11.23 (Dnipropetrovsk) and 7.33 (Kyiv city) to 2.00 (Ternopil) and 2.02 (Rivne).

**Figure 1.**
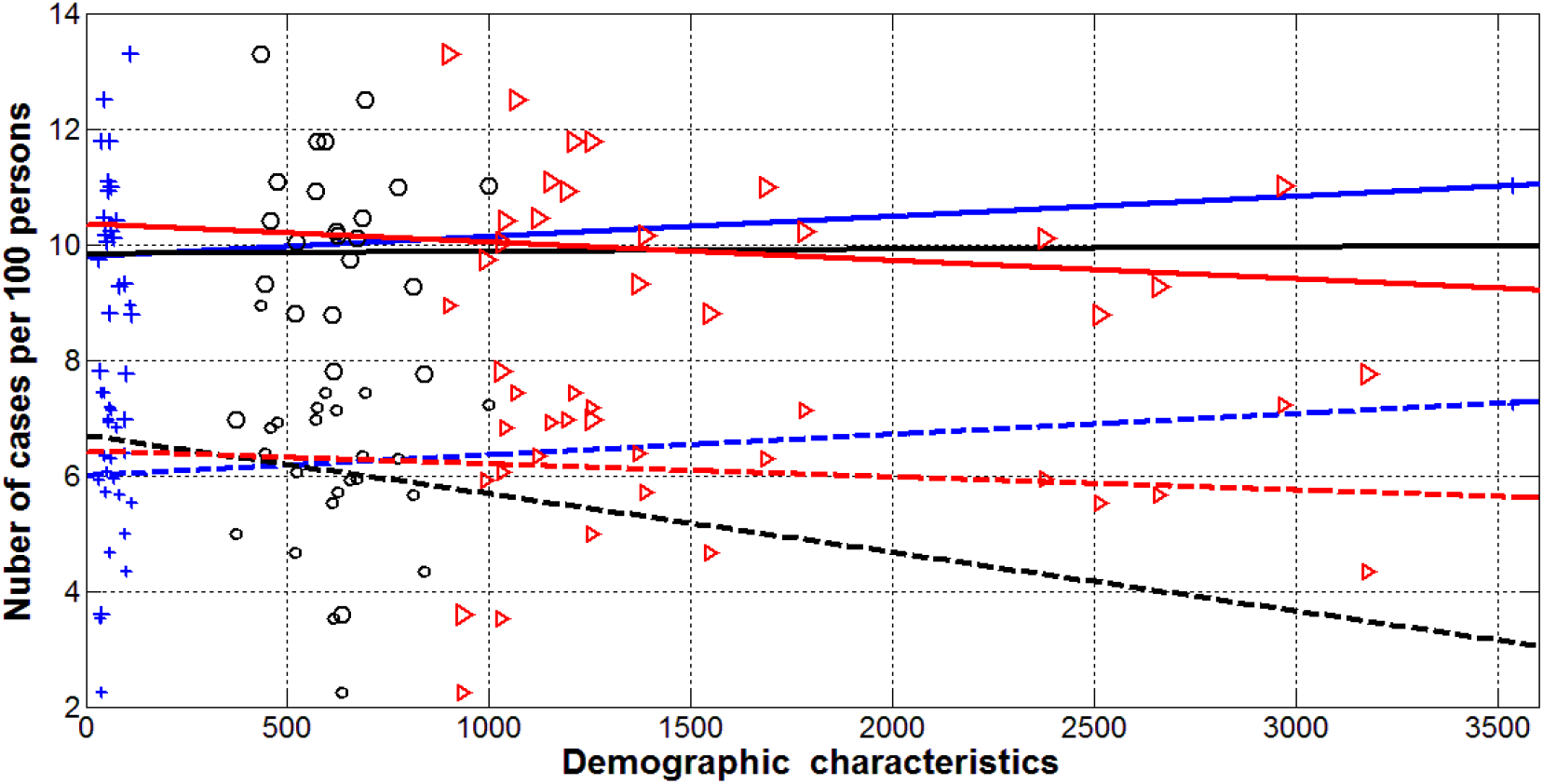
Numbers of COVID-19 cases per 100 persons (CC) in the regions of Ukraine accumulated as of June 27, 2021 versus the volume of population *N*_*pop*_/1000 (red), its density per square km (blue), and the urbanization level 1000\**N*_*urb*_*/N*_*pop*_ (black). Markers represent values from Table 1. Best fitting lines (1) correspond to values shown in Table 3.

As of June 28, 2021, the highest CC levels were registered in Andorra - 18%, Montenegro – 16%, Czech Republic - 15.5%, San Marino - 15%, Slovenia - 12.4%. In six months, the list of the most infected countries has changed as follows: Andorra (27.9%), Montenegro (25.8%), Slovakia (24.6 %). The lowest CC values were in the Vatican (3.3% no new cases were registered last 6 month), Finland (4.1%) and Iceland (6.4%), see Table 2. As of December 23, 2021 the maximum values of deaths per 100,000 persons were registered in Bulgaria (441.3), Bosnia and Herzegovina (405.8) and Hungary (397.6). The minimum values (corresponding to Iceland (10.8), Norway (23.0) and Finland (27.1)) were much smaller, see Table 2. The calculated numbers of deaths per 100 registered cases vary from 4.6 (Bosnia and Herzegovina), 4.2 (Bulgaria) and 3.5 (North Macedonia) to 0.47 (Denmark), 0.34 (Norway) and 0.17 (Iceland).

**Table 3.** Optimal values of parameters in eq. (1), correlation coefficients and the results of Fisher test applications. The results of calculations performed in [20] with the use of data sets corresponding to the end of June 2021 are shown in brackets.

| Number of Figure, relationship, region | $n$ | Correlation coefficient, $r$ | Optimal values of parameters $a$ in eq. (1) | Optimal values of parameters $b$ in eq. (1) | Experimental value of the Fisher function $F$, eq. (2), $m=2$ | Critical value of Fisher function $F_c(1, n-2)$ for the confidence level 0.1, [27] |
| --- | --- | --- | --- | --- | --- | --- |
| 1, CC versus volume of population, Ukraine | 23 | -0.1078<br>(-0.1082) | 10.3503<br>(6.4203) | -3.1785e-007<br>(-2.2551e-007) | 0.2467<br>(0.2486) | 2.96 |
| 2, CC versus volume of population, Europe | 44 | -0.1778<br>(-0.1607) | 14.1528<br>(8.2330) | -4.4192e-008<br>(-2.5094e-008) | 1.3706<br>(1.1132) | 2.84 |
| 1, CC versus density of population, Ukraine | 23 | 0.1232<br>(0.1792) | 9.7826<br>(5.9935) | 3.4716e-004<br>(3.6060e-004) | 0.3235<br>(0.6967) | 2.96 |
| 2, CC versus density of population, Europe | 44 | -0.2026<br>(-0.0953) | 14.1058<br>(8.0331) | -0.0026<br>(-7.7955e-004) | 1.7984<br>(0.3848) | 2.84 |
| 1, CC versus urbanization level, Ukraine | 23 | 0.0028<br>(-0.1024) | 9.8339<br>(6.6956) | 0.0388<br>(-1.0123) | 1.6121e-004<br>(0.2224) | 2.96 |
| 2, CC versus urbanization level, Europe | 44 | -0.1017<br>(-0.0019) | 15.9213<br>(7.8739) | -3.3644<br>(-0.0394) | 0.4387<br>(1.5088e-04) | 2.84 |

Such huge differences raise questions about their cause. As in [20], we have investigated possible correlations with the volume of population, its density and the level of urbanization. The results are presented in Tables 3 and 4 and in Figs. 1-4. The linear regression (1) was used to calculate the regression coefficient *r*, the coefficients *a* and *b* of corresponding straight lines, and the experimental values of the Fisher function. The results are shown in Table 3 for CC values and in Table 4 for DC and 100000*DC/CC values.

**Table 4.**
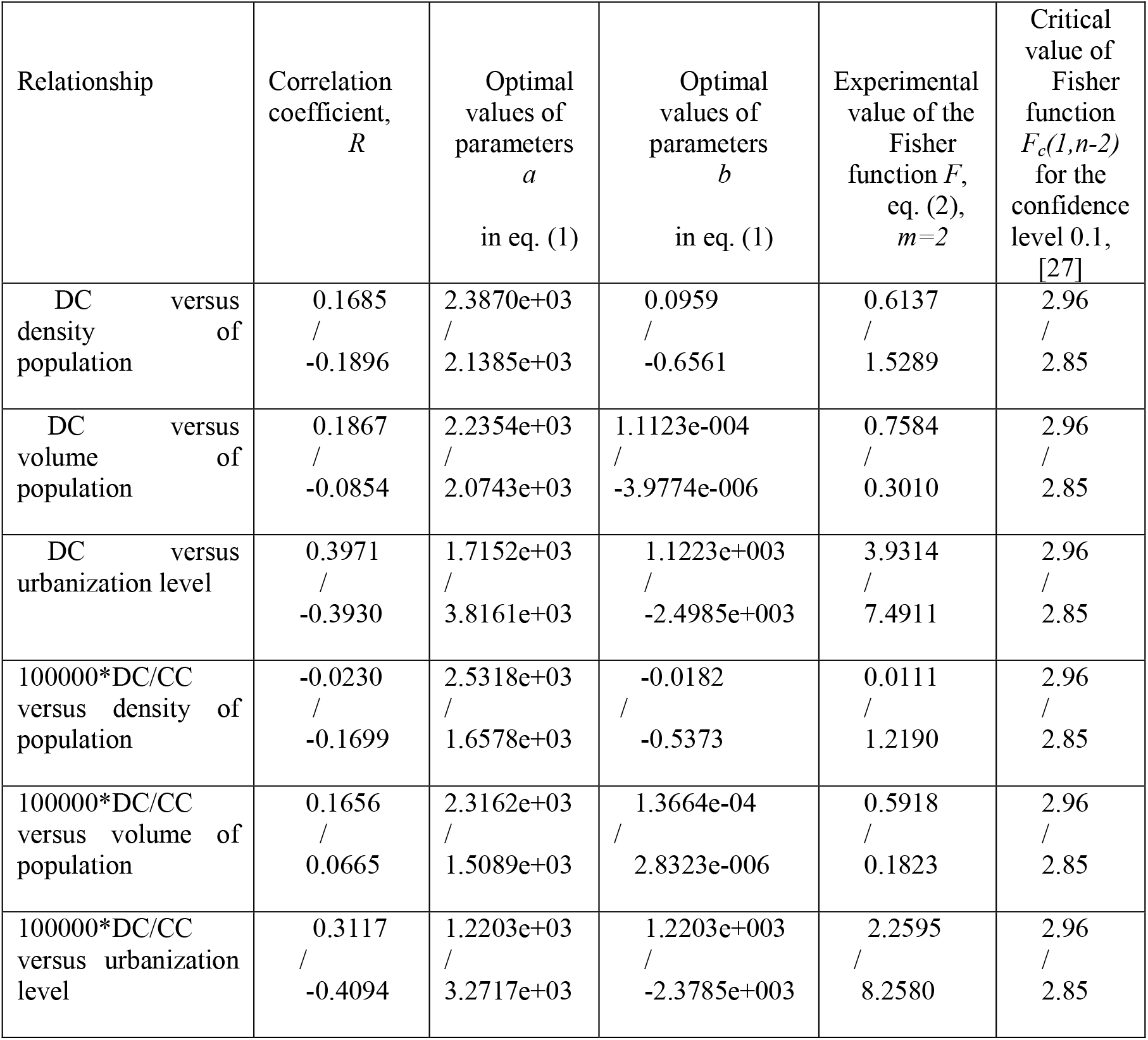
Correlations for ratios of deaths per million (DC) and deaths per 100,000 cases (100000*DC/CC). Optimal values of parameters in eq. (5), correlation coefficients and the results of Fisher test applications. Results for Ukraine are shown before slash, for Europe-after slash. Number of countries in Europe taken for calculations *n*=43 (without Vatican, see Table 2). Number of Ukrainian regions taken for calculations *n*=23 (see Table 1).

The regression analysis demonstrates that there are no correlations between CC values and demographic factors, since *F*_*C*_ (*k*_1_, *k*_2_) > *F* for all cases presented in Table 3 (compare figures in two last columns). This fact is valid both in the case of Ukrainian regions and European countries for datasets corresponding December 23, 2021 and previous study with the use of CC values accumulated at the end of June 2021 (results are published in [20] and shown in brackets). Numbers of cases per 100 persons and corresponding best fitting lines are shown in Fig. 1 for Ukrainian regions and in Fig. 2 for Europe. Small markers and dashed lines correspond to the situation at the end of June 2021, large markers and solid lines – on December 23, 2021. It can be seen that CC data are very scattered. Some visible trends occurred only for dependence CC versus density of population in Europe at the end of 2021 (see the blue solid line in Fig. 2), but even in this case no significant correlation was revealed.

**Figure 2.**
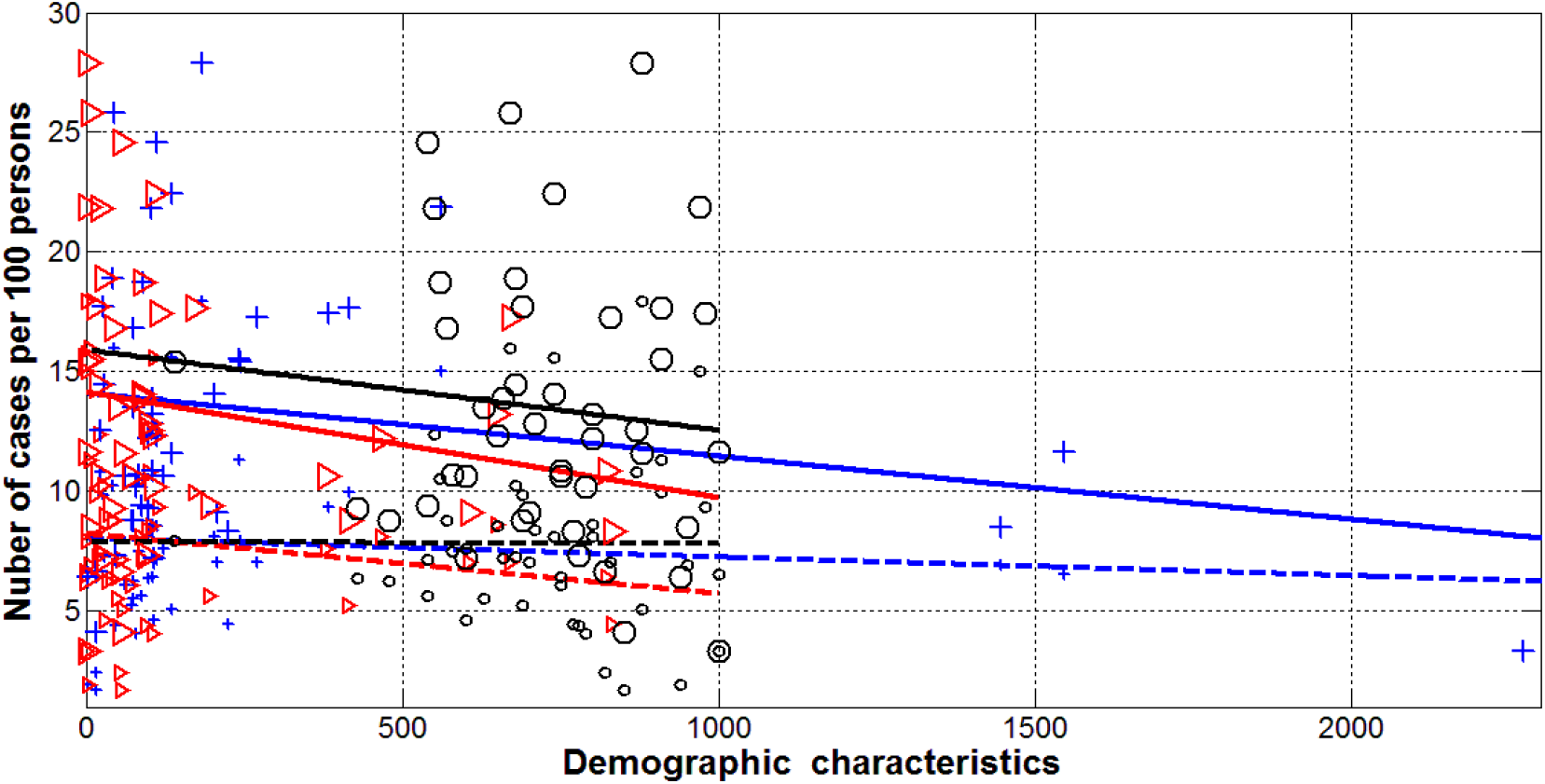
Numbers of COVID-19 cases per 100 persons (CC) in European countries accumulated as of June 28, 2021 versus the volume of population *N*_*pop*_*/100000* (red), its density per square km (blue), and the urbanization level 1000\**N*_*urb*_*/N*_*pop*_ (black). Markers represent values from Table 2. Best fitting lines (1) correspond to values shown in Table 3

The regression analysis of DC values showed that there are correlations with the urbanization levels both for Ukrainian regions and European countries, since *F*_*C*_ (*k*_1_, *k*_2_) < *F* for these cases (see Table 4). The signs of correlation coefficients and parameters *b* are opposite for Ukrainian regions and European countries. In Ukrainian regions the number of deaths increases with the increase of urbanization level (see the black solid line in Fig. 3). The same line in Fig.4 illustrates the opposite trend for European countries. The mortality rate (DC/CC ratio) diminishes with the increase of the urbanization level in Europe (see last row in Table 4 and the dashed black line in Fig. 4). Opposite trend is visible in Fig. 3 for Ukrainian regions, but no statistically significant relationship was revealed (*F*_*C*_ (*k*_1_, *k*_2_) > *F*). The volume of population and its density do not affect the DC and DC/CC values (see Table 4 and Figs. 3 and 4).

**Figure 3.**
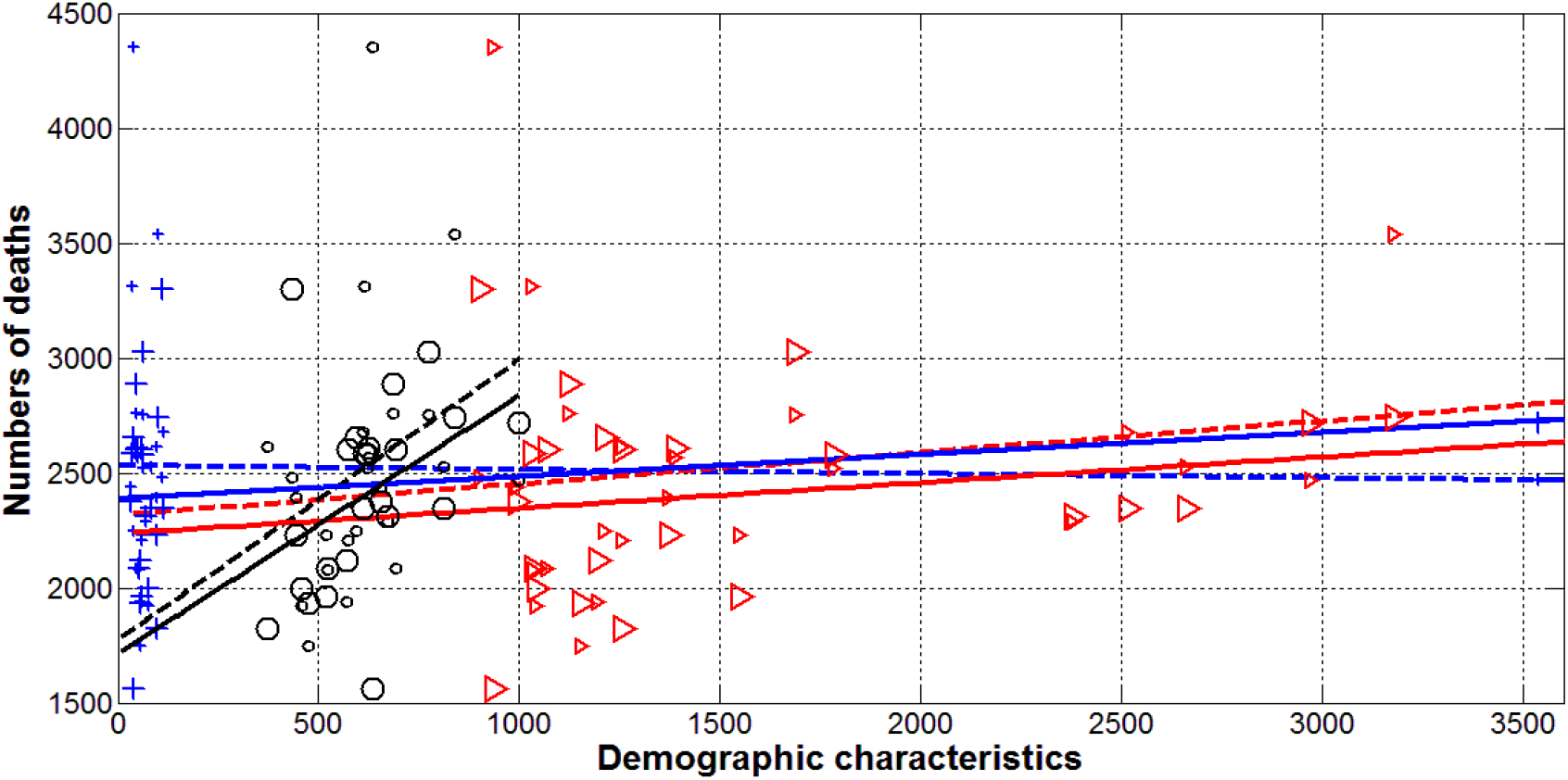
Deaths caused by coronavirus in Ukrainian regions accumulated as of December 23, 2021. Numbers of deaths per million DC (large markers and solid lines) and per 100,000 cases 100000*DC/CC (small markers and dashed lines) versus the volume of population *N*_*pop*_*/1000* (red), its density per square km (blue), and the urbanization level 1000\**N*_*urb*_*/N*_*pop*_ (black). Markers represent values from Table 1. Best fitting lines (1) correspond to values shown in Table 4.

**Figure 4.**
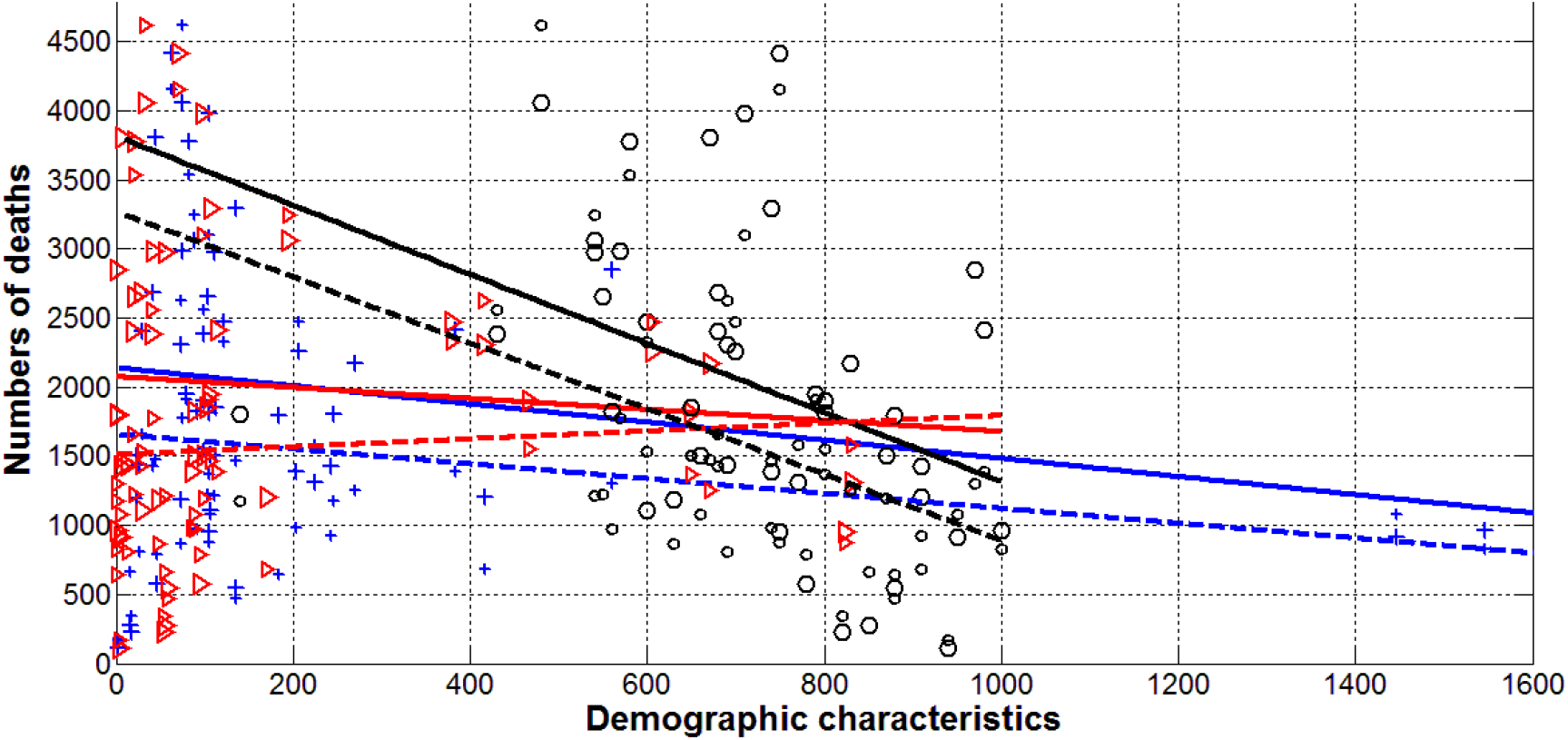
Deaths caused by coronavirus in European countries accumulated as of December 23, 2021. Numbers of deaths per million DC (large markers and solid lines) and per 100,000 cases 100000*DC/CC (small markers and dashed lines) versus the volume of population *N*_*pop*_*/100000* (red), its density per square km (blue), and the urbanization level 1000\**N*_*urb*_*/N*_*pop*_ (black). Markers represent values from Table 2. Best fitting lines (1) correspond to values shown in Table 4.

## Discussion

Very different CC, DC and DC/CC values registered in the regions of Ukraine and European countries do not depend on the volume of population and its density. The deaths per capita and death per case values decrease with the increase of the urbanization level of European countries. Opposite trend was revealed for Ukrainian regions. These facts motivate us to focus on the analysis of quarantine measures, testing, tracing and isolating patients.

We also have to take into account the large number of unregistered cases observed in many countries [28-33]. Estimates for Ukraine made in [28, 33] showed that the real number of cases could be 4-20 times higher than registered and reflected in the official statistics. Probably, in Ukrainian villages many deaths caused by coronavirus were not registered. That is why the DC values increase with the urbanization level in Ukrainian regions. The decrease of DC and DC/CC values in European cities probably are connected with better testing, isolation, and treating the COVID-19 patients.

The results of this study motivate us to pay attention to the pollution effects. The smallest CC, DC, and DC/CC values were registered in the cleanest North European countries (Iceland, Norway, Denmark, and Finland). Higher figures for Sweden are probably connected with the absence of lockdown in 2020. The increase of DC values in the most urbanized Ukrainian regions may be also connected with the atmospheric pollution.

## Data Availability

All data produced in the present work are contained in the manuscript

